# Subtypes of Older Adults Starting Long-Term Care in Japan: Unsupervised Machine Learning Approach

**DOI:** 10.1101/2024.12.24.24319593

**Authors:** Yoko Hamasaki, Masao Iwagami, Jun Komiyama, Yuji Ito, Yuta Taniguchi, Ryota Inokuchi, Taeko Watanabe, Tadahiro Goto, Naoaki Kuroda, Ai Suzuki, Satoru Yoshie, Tatsuro Ishizaki, Katsuya Iijima, Nanako Tamiya

## Abstract

**Objectives:** The older population requiring long-term care (LTC) exhibits heterogeneity in physical and cognitive functions; however, an established classification is lacking. We aimed to identify distinct subgroups of older adults with LTC needs using unsupervised machine learning and to examine differences in their prognoses.

**Design:** Retrospective cohort study.

**Setting and participants:** Using survey data for care-need certification (linked to LTC and medical insurance claims) in City A, Japan, we identified community-dwelling adults aged ≥ 65 years who started LTC. Data from City B were used for validation of clustering.

**Methods:** We applied latent class analysis to group the participants in City A, based on all 74 items (38 on physical functions, 9 on cognitive functions, 15 on behavioral problems, and 12 on medical procedures) in the Japanese standardized care-needs certification survey. Then, we examined the association between the identified subtypes and four outcomes, including death, hospitalization, nursing home admission, and care-need level deterioration, using regression models.

**Results:** Among 3,841 participants in City A (median age, 83 years; 59.3% female), five subtypes were identified: (i) mild physical, (ii) mild cognitive, (iii) moderate physical, (iv) moderate multicomponent, and (v) severe multicomponent. The results of clustering were replicated in City B. Compared with the mild physical subtype, the severe multicomponent subtype showed the highest risk of death (adjusted hazard ratio [aHR] 2.56; 95% confidence interval [CI] 2.02–3.24), and nursing home admission (aHR 5.91; 95% CI 4.57–7.63). The moderate physical subtype showed a higher risk of hospitalization (aHR 1.32; 95% CI 1.16–1.49), and the moderate multicomponent subtype was more likely to experience care-need deterioration (adjusted odds ratio 1.67; 95% CI 1.26–2.22).

**Conclusions and Implications:** This study identified five subtypes of older adults who started LTC. These findings inform individualized care decisions and tailored planning of medical and LTC services.

## Introduction

The number of older individuals with disabilities due to aging or disease is increasing globally.^1^ Many of these individuals have coexisting diseases and disabilities,^2^ presenting complex healthcare challenges.^3–5^ Thus, managing a single disability or disease is often insufficient, necessitating tailored care strategies.^6^ Identifying subgroups of older adults with disabilities and understanding their prognoses can help healthcare providers develop more tailored care plans. However, to our knowledge, there has been no classification system for older adults requiring long-term care (LTC) based on their physical and cognitive (behavioral) functions.

Japan, which has the world’s highest proportion of older adults, established a long-term care insurance (LTCI) system in 2000 to support older people regardless of their income level or informal care availability.^7,8^ Under the LTCI system, eligibility for LTC services is measured as a care-need level (7-level ordinal scale) based on the estimated time required for care evaluated by a 74-item survey, which was nationally standardized by a government’s expert committee.^9,10^ However, the care-need level alone is insufficient to understand the complex condition of older adults requiring LTC because this scale is only based on the time needed for LTC services. Therefore, another classification based on the patterns of the 74-items (consisting of objectively measured physical and cognitive functions, behavioral problems, and required medical procedures) and clarifying their prognoses could be useful for considering tailored care.

We aimed to identify unique and relatively homogeneous subgroups of community-dwelling older adults requiring LTC in Japan based on a nationally standardized care-needs certification survey and to examine whether their prognoses (death, hospitalization, nursing home admission, and care-need level deterioration) differ among these subgroups.

## Methods

### Data Sources

We obtained data from care-need certification surveys (February 2012 to March 2019), medical and LTC insurance claims data and insurance registration data (April 2014 to March 2019) in City A, Japan. Care-need certification survey data between February 2012 and September 2014 were used to exclude those with records of certification surveys before October 2014 (Supplementary Figure S1) and identify the first certification. Data were collected by local government and linked using anonymized unique identifiers of the recipients. City A’s population was 240,383, including 46,613 (19.4%) people aged ≥ 65 in 2018.

The care-need certification survey was conducted by certified local government surveyors who assessed LTC applicants in their homes or inpatient facilities.^9^ The survey includes 74 items on physical functions (38 items), cognitive functions (9 items), behavioral problems (15 items), and medical procedures (12 items).^11^ Based on the survey and physician’s opinion letter, each applicant receives LTC need certificates, ranging from support levels 1 and 2 and care need levels 1 (least disabled) to 5 (most disabled). Medical insurance claims data included diagnoses, medical procedures, and prescriptions from inpatient and outpatient care. The LTC insurance claims data included information on the types of services and dates of admission to or discharge from a nursing home.^12^ Insurance registration data captured the dates and reasons for loss of eligibility, such as death. This study was approved by the ethics committee of the University of Tsukuba (approval number: 1445-15 and 1666-1). Informed consent from individuals was waived because of the anonymous nature of the data.

### Study Design and Population

A cross-sectional analysis was conducted for clustering, followed by a retrospective cohort analysis to examine the association between subtypes and prognoses. Supplementary Figure S2 shows the design diagram of the study population, covariates, and outcomes.

We identified individuals aged ≥ 65 years who received the LTC certification survey between October 1, 2014, and March 31, 2019, and were certified at care-need levels ranging from 1–5 for the first time. We did not include those with support levels 1–2 because most of their physical and cognitive functions were not impaired. Participants unable to be matched with claims data were excluded.

### The 74-items in the Japanese Standardized Care-needs Certification Survey

Seventy-four items from the care-needs certification survey at baseline were obtained from the survey data. These include 38 items on physical functions (e.g., walking, eating, shopping), nine items on cognitive functions (e.g., communicate intentions to others, understanding of daily routine, remembering own name), 15 items on behavioral problems (e.g., frequency of feeling persecuted, making up a story, emotional instability), and 12 items on required medical procedures (Table 1). While some items had 3 or more categories (e.g., not assisted, partly assisted, or fully assisted), each variable was converted to a binary form, with 1 if the individual required assistance or a medical procedure, and 0 otherwise.

**Table 1.** Seventy-Four Items and Dimensions of the Survey Data for Care-Need Certification in Japan.

| Dimensions (number of items) | Items |
| --- | --- |
| **Physical function (38)** |  |
| Body function /Bed mobility (20) | paralysis, joint contracture, turning over in bed, sitting up in bed, sitting up, both-leg standing, walking, standing up, single-leg standing, washing the whole body, trimming the nails, eyesight, hearing ability |
| Activities of daily living (12) | transferring, moving, swallowing, eating, regulating urination, regulating defecation, oral hygiene, washing face, combing/styling hair, upper-body dressing, lower-body dressing, frequency of going out |
| Instrumental activities of daily living (6) | taking medicine, managing money, daily decision making, maladaptation to group, shopping, cooking |
| **Cognitive function (9)** | communicate intentions to others, understand daily routine, remember date of birth, short-term memory, remembering own name, recognizing the season, location awareness, wandering, being lost |
| **Behavioral problems (behavioral and psychological symptoms) (15)** | frequency of feeling persecuted, making up a story, emotional instability, reversal of day and night, repeating the same story, shouting, resisting advice or care, restlessness, frequent behavior to go out alone, collecting items inappropriately, destruction of things/ clothes, forgetfulness, monology, selfish behavior inappropriate for the situation, incoherent talk |
| **Medical procedures (12)** | intravenous infusion, intravenous hyperalimentation, dialysis, care for ostomy, oxygen therapy, artificial ventilator, care for tracheostomy, pain care, tube feeding, monitoring (such as blood pressure, heart rate, oxygen saturation), pressure ulcer care, incontinence care (such as condom catheter, indwelling catheter, urostoma) |

### Outcomes

For the longitudinal analysis, we evaluated the four outcomes separately: all-cause mortality, hospitalizations, nursing home admissions, and deterioration of care-need levels. Nursing homes included LTC welfare facilities (for those whose condition is stable but require regular care with care-need level ≥ 3), LTC health facilities (for those requiring care and rehabilitation to return home), integrated facilities for medical and LTC, LTC medical facilities (for individuals requiring substantial care and medical

services), specified facilities(home-like facilities for those whose condition is stable but require LTC), and group homes (home-like facilities for those with mild to moderate dementia). These services are all covered by the LTCI.^8,13,14^ Care-need level deterioration was defined as an increase in care-need level or death within 2 years of the initial survey.

### Statistical Analyses

First, we used latent class analysis (LCA) to identify subpopulations of older adults requiring LTC at baseline (i.e., the first time each participant received the care-needs certification survey between October 2014 and March 2019). LCA is a finite mixture modeling method that assumes the overall population heterogeneity of observable response variables results from unobserved, homogenous subgroups known as latent classes.^15,16^ The posterior probability of belonging to each class was obtained for each participant, and individuals were assigned to the cluster with the highest posterior probability.

LCA models were fitted, starting with a two-subtype model and increasing the number of subtypes. The optimal number of subtypes was determined based on model fit indices, classification accuracy, clinical relevance, and interpretability. Indices included the Bayesian Information Criterion (BIC), Akaike’s Information Criterion (AIC) (smaller values of those indicate better model fit), and elbow method (plot a fit statistic and identify where the fit visually changes).^17^ Classification accuracy was assessed by the average posterior probability of subtype membership, calculated as the mean of the members’ posterior probabilities, with 0.8 suggesting clear classification.^18^

Subtypes were labeled (i.e., named) based on the 74 items that distinguished them. Observed/expected (O/E) ratios and exclusivity measures characterized the subtypes. The O/E ratio was calculated by dividing the proportion of each item in a cluster by the proportion in the total study population. Exclusivity was calculated as the proportion of the number of participants with an item in a

cluster to the total number of participants with that item. The items were identified as components of specific subtypes when the O/E ratio was ≥ 2, and exclusivity was ≥ 25%.^19,20^

In the longitudinal analyses, we estimated survival curves using the Kaplan–Meier method, followed by multivariate Cox regression analysis to estimate adjusted hazard ratios (aHRs) for death, adjusting for age and sex. The population was followed up until either death or loss to follow-up due to exclusion from the insurance registry for reasons such as moving, or until March 31, 2019 (end of the study period).

For hospitalizations and nursing home admissions, we used cumulative incidence functions that accounted for competing risks of death to estimate cumulative incidence, and Fine–Gray regression models^21,22^ to estimate sub-distribution hazard ratios, adjusting for age and sex. The population was followed up until either the first record of the outcome, death, loss to follow-up, or March 31, 2019, whichever occurred first. For nursing home admissions, those already admitted to nursing homes at baseline were excluded.

Care-need level deterioration within 2 years was compared among subtypes, limited to participants who started care before April 2017 to ensure all participants (except for those who became lost to follow-up) had two years of follow-up. The care needs status (improved, no change, worsened, died, not reassessed, or lost to follow-up) was described 2 years after the certification survey. Univariate and multivariable logistic regression analyses adjusted for age and sex were performed to address the worsening care needs. For those who were lost to follow-up and not reassessed, the care-need level was assumed to remain unchanged. For the sensitivity analysis, we excluded those who were lost to follow-up and not reassessed.

For death, hospitalizations, and nursing home admissions, we also conducted stratified analyses by care-need level 1–2 or 3–5.

The level of significance was set at P <.05 (two-tailed). All LCA models were estimated using the poLCA package in R version 3.60 (Foundation for Statistical Computing, Vienna, Austria). Stata/MP version 17 (StataCorp, College Station, TX, USA) was used for all other statistical analyses.

### External Validation

To evaluate the external validity of our clustering approach, we replicated the analysis using data in City B, Japan, with a population of 48,444, of whom 17,329 (35.8%) were aged ≥ 65 years. Using City B’s care-need certification survey data (April 2012 to March 2014), we identified the participants between October 2012 and March 2014 with the same inclusion and exclusion criteria. The same clustering procedure was repeated and compared with the results from City A. Only cross-sectional analyses were performed in City B owing to insufficient follow-up data.

### Additional Analysis

In the main analysis, in line with this study’s purpose to classify older adults starting LTC based on their functions according to the care-needs certification survey, we did not include their underlying diseases, which are considered upstream of their functions. As an additional analysis, we included 22 diseases (presence or absence of each disease) potentially associated with initiating LTC.^23^ We defined each disease using relevant International Classification of Diseases, 10th Revision codes^23^ recorded in the past six months before the care-need certification survey date.

## Results

A total of 3,841 individuals from City A were included (Figure 1). The median age was 83 (interquartile range [IQR], 77–87) years, and 59.3% were females.

**Figure 1.**
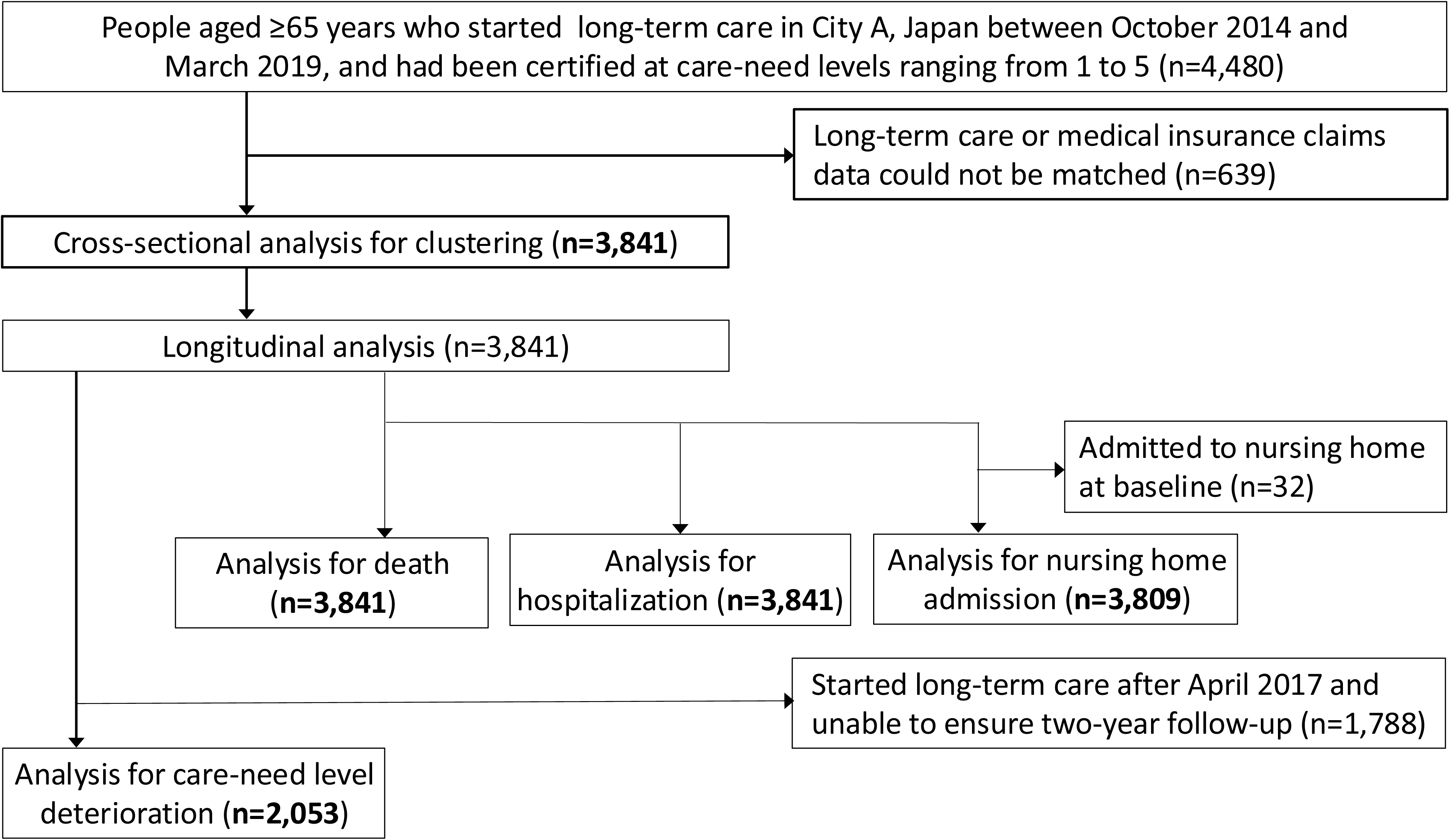
Flow Chart for Study Participant Selection

### Subtypes of Older Adults Starting LTC Based on Clustering Approach

At baseline, up to a nine-subtype model was estimated, and a five-subtype was chosen as the final model. The five-subtype model was considered an optimal fit because the elbow plot showed an "elbow" at 5, further increasing the number of subtypes not yielding the same decrease in AIC and BIC (Figure S3). The model offered the most reasonable clinical interpretability and fulfilled the predetermined minimum required average posterior probability of class membership (0.94) (Table S1).

We labeled the clusters as: (i) mild physical (n = 1258, 32.8%), (ii) mild cognitive (n = 946, 24.6%), (iii) moderate physical (n = 767, 20.0%), (iv) moderate multicomponent (n = 597, 15.5%), and (v) severe multicomponent subtypes (n = 273, 7.1%) (Figure 2). Detailed characteristics of each subtype are presented in the Supplementary Tables S2 and S3. Age distribution was similar across subtypes, whereas the mild cognitive subtype had the highest proportion of females (66.1%) (Table 2). The mild physical and mild cognitive subtypes tended to include older adults with milder care-need levels, whereas the moderate physical and severe multicomponent subtypes tended to include those with more severe care-need levels.

**Figure 2.**
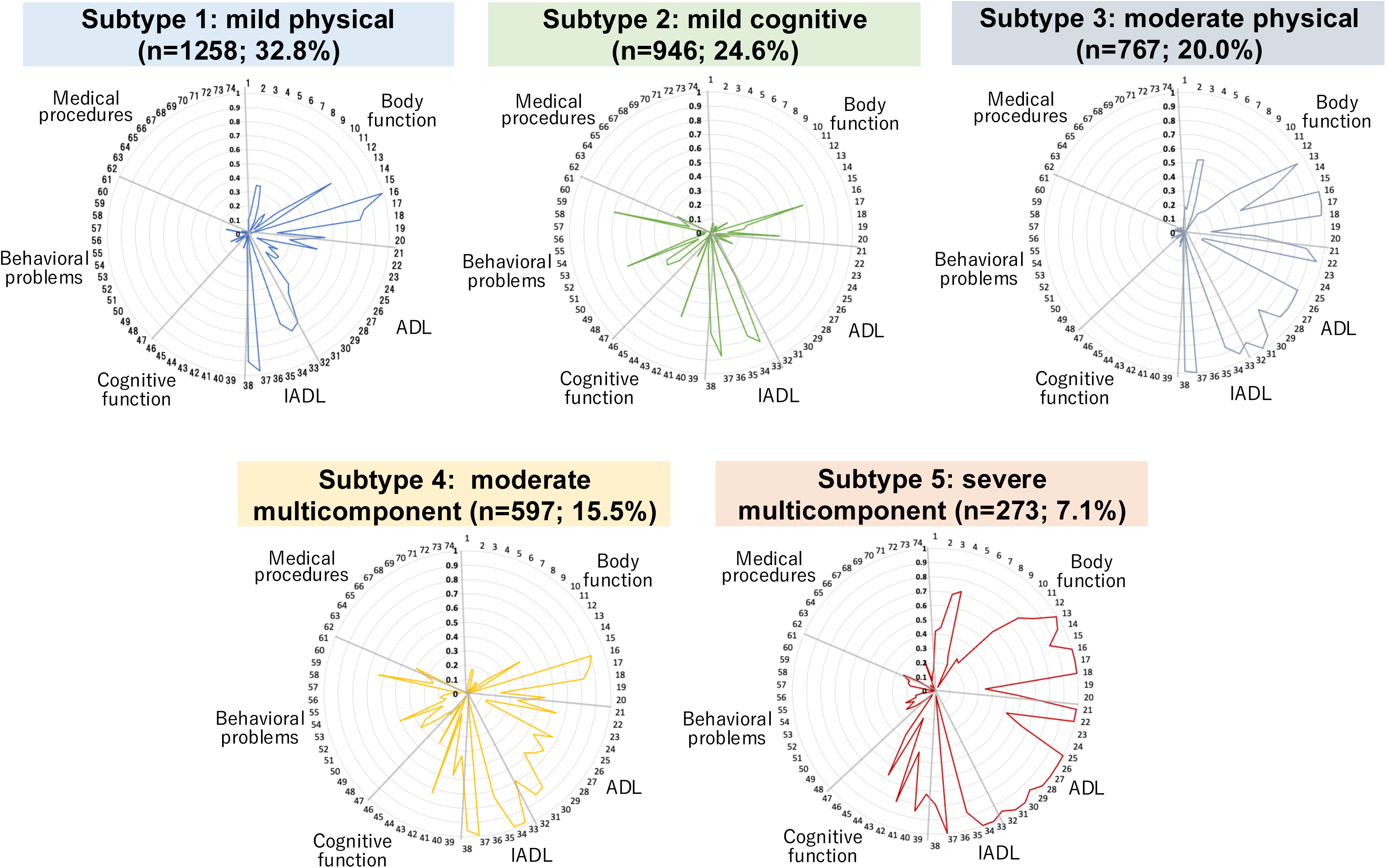
Subtypes in Older Populations Starting Long-Term Care in Japan. Abbreviation: ADL, activities of daily living; IADL, instrumental activities of daily living. The figure shows the item response probabilities (ranging from 0–1) across the subtypes, with each number corresponding to the care needs certification survey items in Table S2.

**Table 2.**
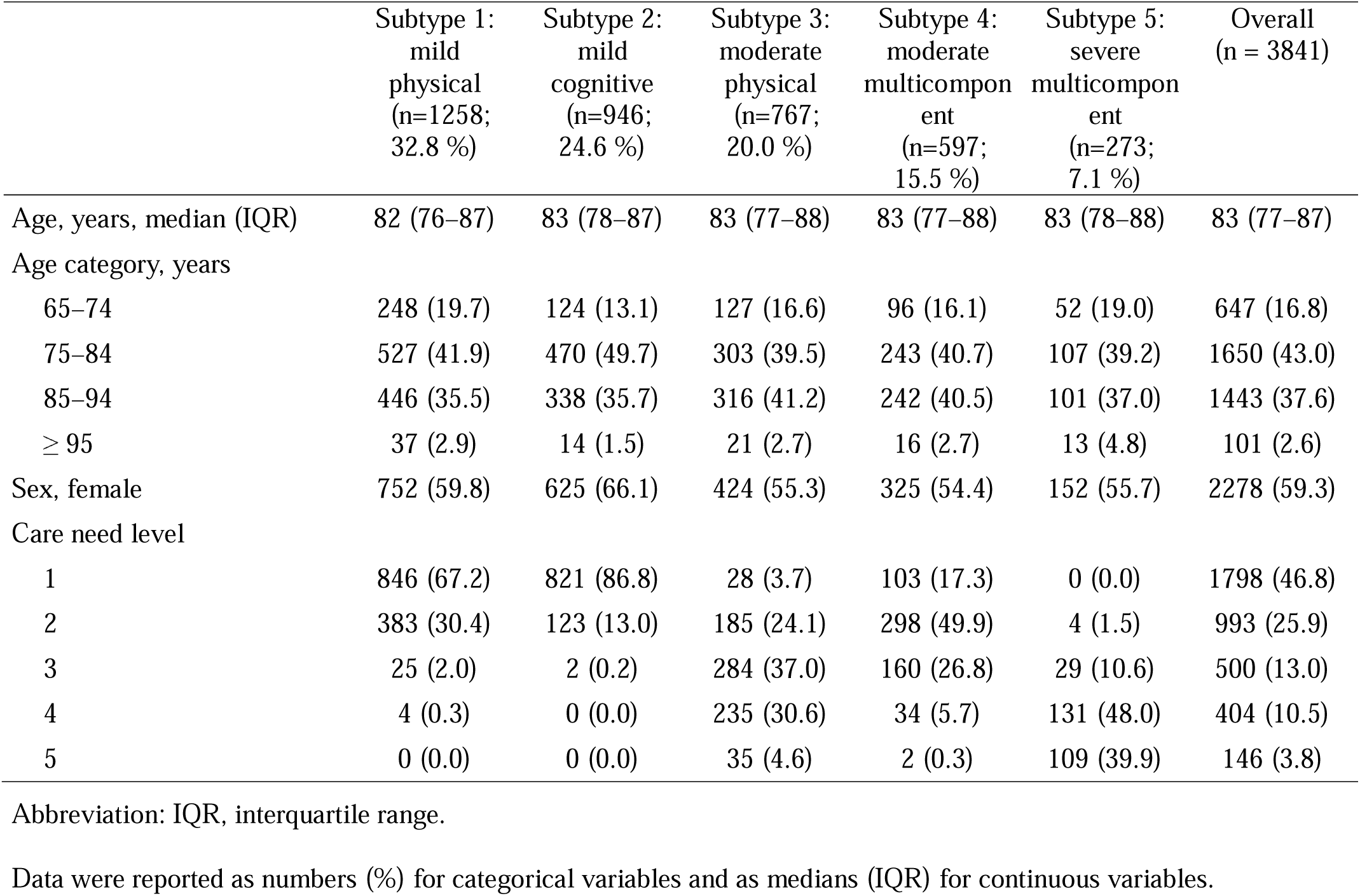
Baseline Characteristics by Physical and Cognitive Functions, Behavioral Problems, and Medical Procedure Subtypes.

For the external validation using the City B data (n = 2911), the distribution of subtypes and baseline demographic patterns were similar to those of City A (Figure S4 and Table S4). In the analysis which additionally included 22 diseases in LCA, the optimal number of subtypes was similarly considered 5. The distribution of the 74-items in each subtype was similar to that of the main analysis (Table S5). The mild physical and moderate physical subtypes had a high prevalence of ischemic heart diseases, cancer, joint diseases, and fractures. In contrast, cerebrovascular diseases and pneumonia were most prevalent in the severe multicomponent subtype. The mild cognitive and moderate multicomponent subtypes had a high prevalence of dementia.

### Comparison of Outcomes Between Subtypes

Over the median follow-up of 19.0 (IQR, 9.6–33.5) months, 832 deaths occurred. The incidence of outcomes in each subtype is shown in Tables S6 and S7, and their survival curves and cumulative incidence curves are plotted in Figures S5, S6, and S7. As presented in Figure 3, the severe multicomponent subtype had the higher aHR (95%CI) for death (2.56 [2.02–3.24]), hospitalization (1.23, [1.02-1.48]) and nursing home admission (5.91 [4.57–7.63]) compared with the mild physical subtype. The moderate physical subtype had a higher aHR (95%CI) for hospitalization (1.32, [1.16–1.49]), and the moderate multicomponent subtype had an increased risk of care-need level deterioration (adjusted odds ratio, 1.67; 95%CI 1.26–2.22). The mild cognitive subtype had lower aHRs (95%CI) for mortality (0.72 [0.58–0.89]) and hospitalization (0.74 [0.65–0.83]) compared with the mild physical subtype. Results of multivariable analyses are also shown in Tables S6 and S7. Sensitivity analysis for care-need level deterioration yielded similar findings. The association between subtype and outcomes were similar in subgroups with care-need level 1—2 and 3—5 (Table S8).

**Figure 3.**
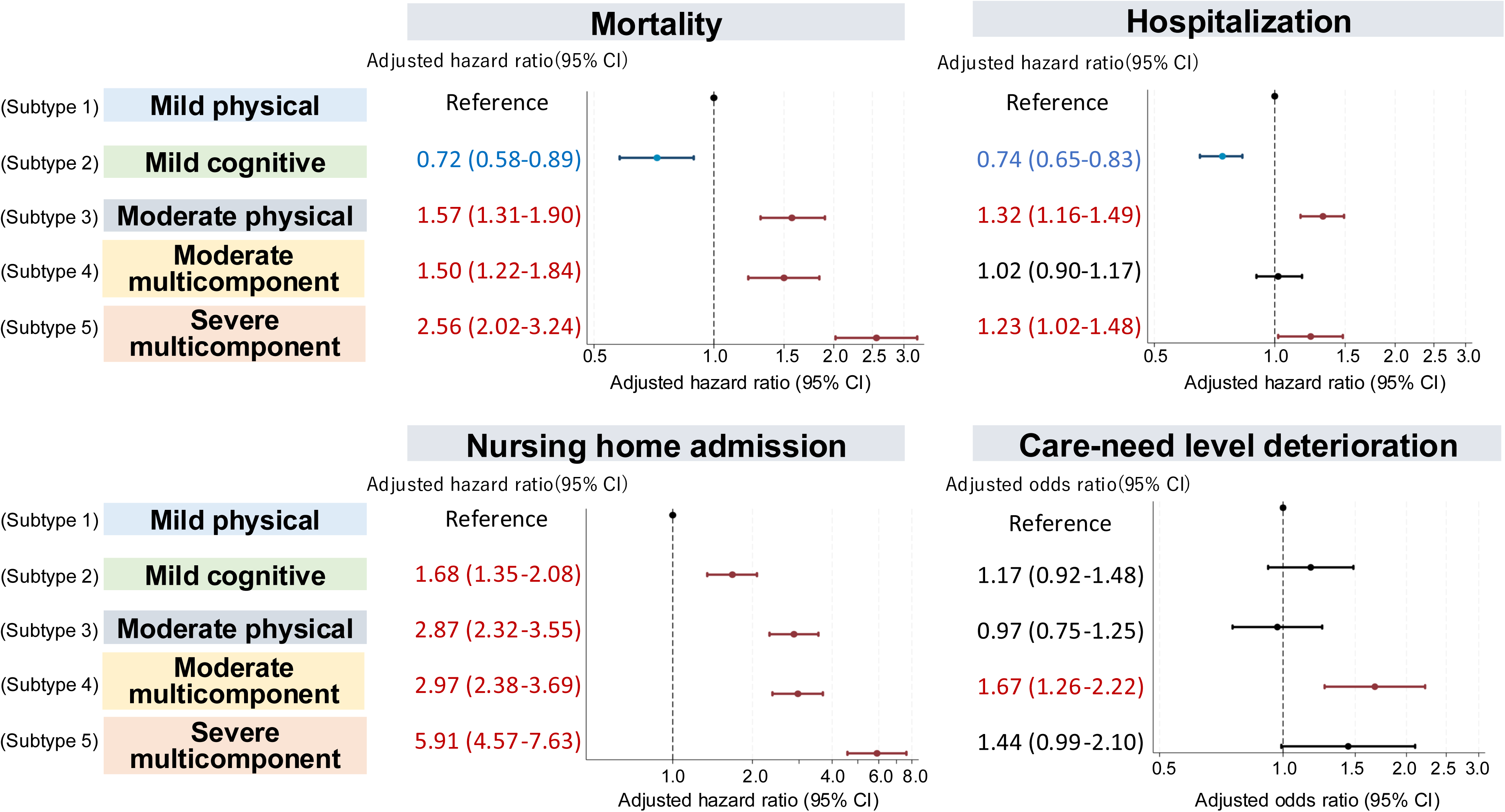
Association Between Subtypes of Physical and Cognitive Functions, Medical Procedures, and the Outcomes **Abbreviation: CI, confidence interval. All the regression models were adjusted for age and sex.**

## Discussion

### Summary of Findings

We identified five subgroups after subtyping 3,841 older adults certified for LTC, based on a nationally standardized care-needs certification survey in Japan. Prognoses varied significantly between the subtypes. The mild physical subtype had mild physical impairments (e.g., standing and moving) but maintained cognitive functions. The mild cognitive subtype had almost no impairment in physical function but only mild cognitive decline with behavioral and psychological symptoms in dementia (BPSD) and had a lower risk of death and hospitalization but a higher risk of nursing home admission than the mild physical subtype. The moderate physical subtype exhibited moderate physical impairment (excluding swallowing and eating) but little cognitive impairment, with higher risks of death, hospitalization, and nursing home admission. The moderate multicomponent subtype had moderate physical and cognitive impairment, with about half experiencing communication difficulties. This subtype had higher risks of death, care needs worsening, and nursing home admission. The severe multicomponent subtype involved severe physical and cognitive impairment, with over 70% having impaired communication. This subtype had a higher risk of death, hospitalization, and nursing home admission.

### Comparison with Previous Studies

One study from Australia using data-driven clustering approaches to subtype older people (≥ 85 years) revealed four subtypes based on the number of diseases, presence of mental health conditions, and activities of daily living (ADL)/instrument ADL scores.^24^ The study reported the subgroup with depression and dementia had a higher mortality. Unlike previous studies, we used detailed information on physical and cognitive functions and medical procedures measured by trained experts. We found a higher risk of death in the moderate and severe multicomponent subtypes, which had a higher proportion of cognitive impairment, similar to previous studies. Unique to our study was the mild cognitive subtype, which had a lower risk of mortality and hospitalization despite cognitive impairment. Furthermore, our study showed a wide range of prognoses varied by these five subtypes, whereas the previous study only evaluated the association with mortality.

### Possible Mechanisms

The higher mortality risk in the moderate physical, moderate multicomponent, and severe multicomponent subtypes may be explained by the differences in physical and cognitive functions^25^ or underlying diseases such as cancer, cardiac disease, pneumonia, and cerebrovascular disease, which are the leading causes of death among Japanese older adults.^26^ For example, the severe multicomponent subtype had a higher prevalence of cerebrovascular disease and physical and cognitive impairment, which may be linked to the highest risk of death. The results suggest the importance of assessing not only diseases but also functional aspects. Moreover, for subtypes with a particularly high mortality risk, early implementation of advanced care planning may be worthwhile.

Regarding hospitalization, the risk was higher for the moderate physical and severe multicomponent subtypes. These subtypes had a wide range of impaired ADLs and diseases that could exacerbate their condition, such as ischemic heart disease and pneumonia. The coexistence of these impairments and diseases may indicate a higher need for medical management, leading to hospitalization. For these subtypes, home visits by healthcare professionals may help manage their conditions and avoid hospitalization.

For nursing home admissions, all other subtypes were at higher risk than the mild physical subtype, consistent with previous studies reporting cognitive and physical impairments are associated with institutionalization.^27^ The present study showed that even the mild cognitive subtype had a higher risk of nursing home admission. The reason for nursing home admission is unclear, but cognitive decline may have increased the caregiver’s burden, as BPSD is reportedly associated with caregiver burden.^28^ Our findings indicate that even when cognitive decline is mild, it is important to assess whether there are factors that make living at home difficult, such as anxiety about their own lives and the burden of care on family caregivers.

Regarding the deterioration of care-need levels, only the moderate multicomponent subtype had a worse outcome. A previous study showed those with intermediate cognitive impairment tended to feel uncomfortable asking doctor questions and to avoid doctors owing to embarrassment, which may be linked to less meaningful engagement with healthcare.^29^ Thus, the higher risk for worsening their condition among the moderate multicomponent subtype may be explained by the cognitive impairment that could affect the timely detection and appropriate management of their conditions.

### Limitations

This study has several limitations. First, this study was conducted in one Japanese city, and thus the generalizability may be limited. Although clustering results were validated using data from another city, the external validity should be critically evaluated when applying the findings to other cities and countries. Second, like any study using clustering methods, labeling each subtype could be subjective. Different researchers can name each subtype differently based on their knowledge and experience.

### Conclusions and Implications

We identified five subtypes of older adults starting LTC in Japan, each with varying physical and cognitive functions, behavioral problems, and medical procedures. Their prognoses varied among the subtypes. These subtypes provide valuable insights for tailoring individualized care for older populations requiring LTC.

## Data availability

The data that support the findings of this study are available from City A and City B, Japan, but restrictions apply to the availability of these data, which were used under license for the current study, and so are not publicly available. Data are however available from the authors upon reasonable request and with permission of City A and City B.

## Brief Summary

This study identified five subtypes of older adults starting long-term care in Japan, each with unique profiles. These findings highlight the need for personalized care strategies.

## Acknowledgments

We thank Editage (http://www.editage.com) for English language editing.

