## Supplementary Materials for "Subtypes of Older Adults Starting Long-Term Care in Japan: Unsupervised Machine Learning Approach"

**Figure S1.** Obtained Survey Data for Care-Need Certification, Medical Claims Data, Long-Term Care Insurance Data, and Insurance Registration Data in City A

**Figure S2.** Study Design

**Figure S3.** Elbow Method for Determining the Optimal Number of Subtypes

**Figure S4.** Proportion of Each Subtype in City A and City B in the Overall Population

**Figure S5.** Kaplan–Meier Survival Curves for Each Subtype

**Figure S6.** Cumulative Incidence of Hospital Admission for Each Subtype

**Figure S7.** Cumulative Incidence of Nursing Home Admission for Each Subtype

**Table S1.** Comparisons of the Model Fit Between the Different Numbers of Subtypes

**Table S2.** Item Response Probabilities for Physical and Cognitive Functions, Behavioral Problems, and Medical Procedures for Each Subtype in City A

**Table S3.** Item Response Probabilities of Clusters of Physical and Cognitive Functions, Behavioral Problems, and Medical Procedures for Each Subtype in City B

**Table S4.** Baseline Characteristics by Physical and Cognitive Functions, Behavioral Problems, and Medical Procedures Subtypes in City B

**Table S5.** Results of Additional Analysis Including Underlying Diseases for Clustering in City A

**Table S6.** Details of the Longitudinal Analysis and Results of the Cox Regression Analysis for Death, Hospitalization, and Nursing Home Admission

**Table S7.** Details of Changes in Care-Need Levels and Results of Logistic Regression Analysis for Care-Need Level Deterioration within 2 Years

**Table S8.** Subgroup Analysis by Care-Need Level: Results of the Cox Regression Analysis for Death, Hospitalization, and Nursing Home Admission

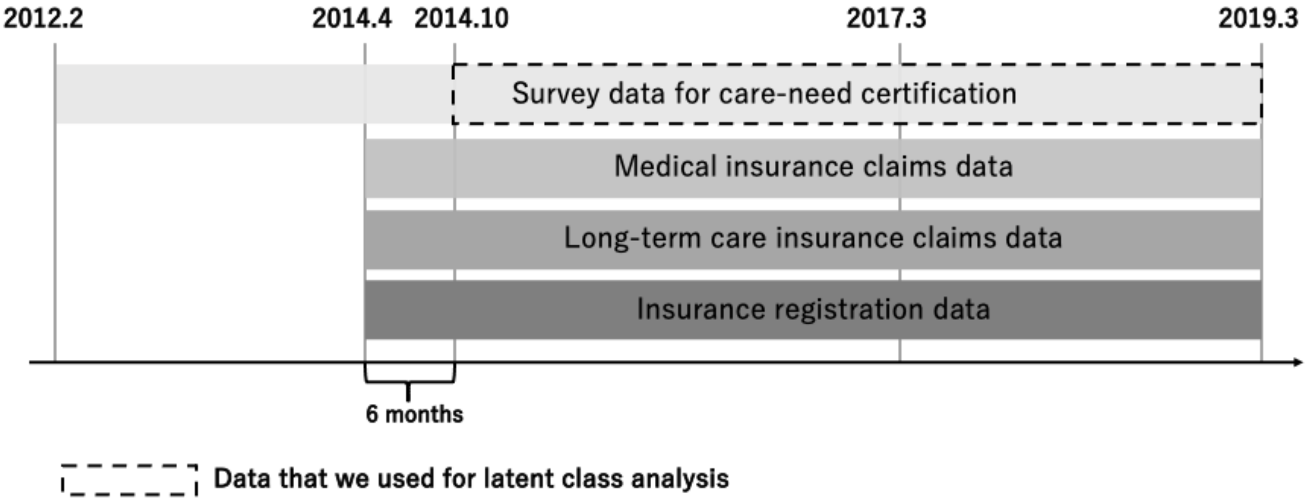


**Figure S1. Obtained Survey Data for Care-Need Certification, Medical Claims Data, Long-Term Care Insurance Data, and Insurance Registration Data in City A**

Survey data for care-need certification between February 2012 and March 2014 were used to identify and exclude those with past records of the certification survey before April 2014. Medical insurance data between April and September 2014 was used to identify baseline comorbidities.

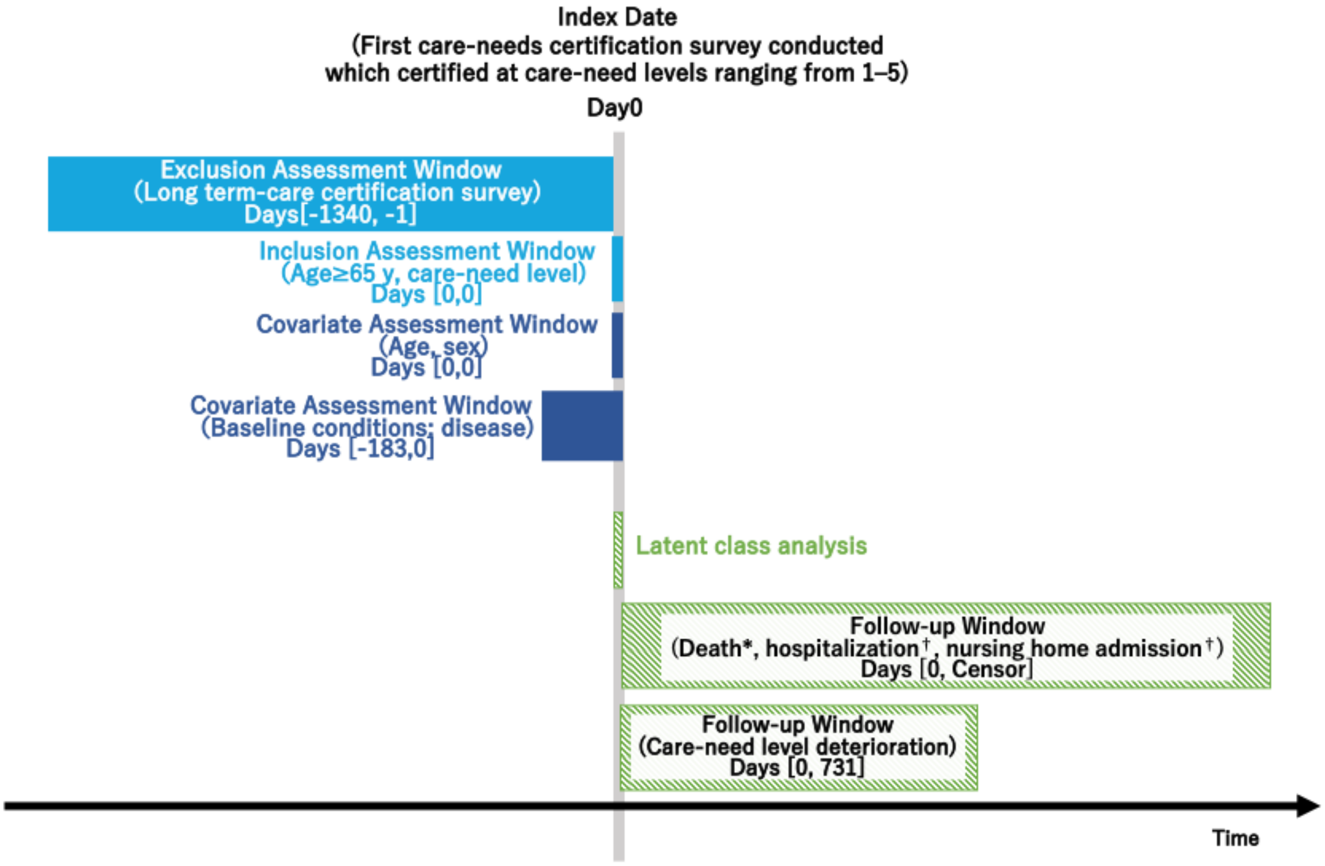


**Figure S2. Study Design**

*Earliest outcome of interest, moving, or end of study period.

†Earliest outcome of interest (hospitalization or nursing home admission), death, moving, or end of study period. Death was treated as a competing risk.

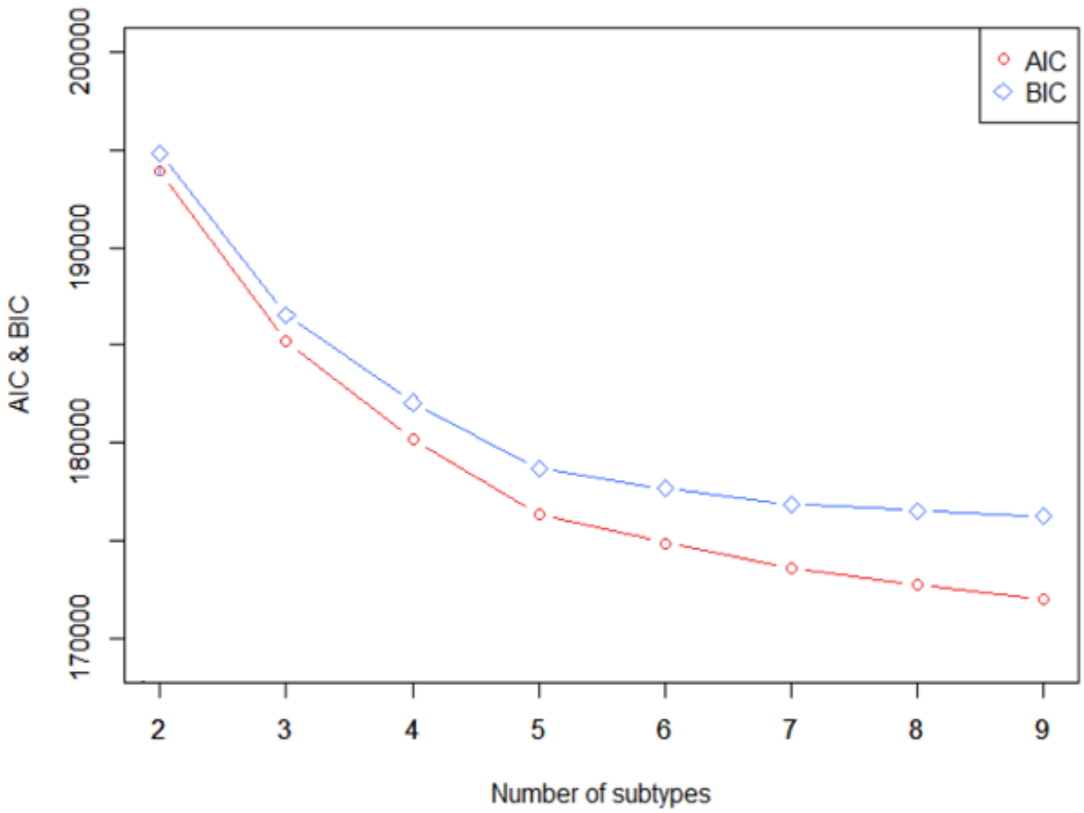


**Figure S3. Elbow Method for Determining the Optimal Number of Subtypes**

Abbreviations: AIC, Akaike Information Criterion; BIC, Bayesian Information Criterion

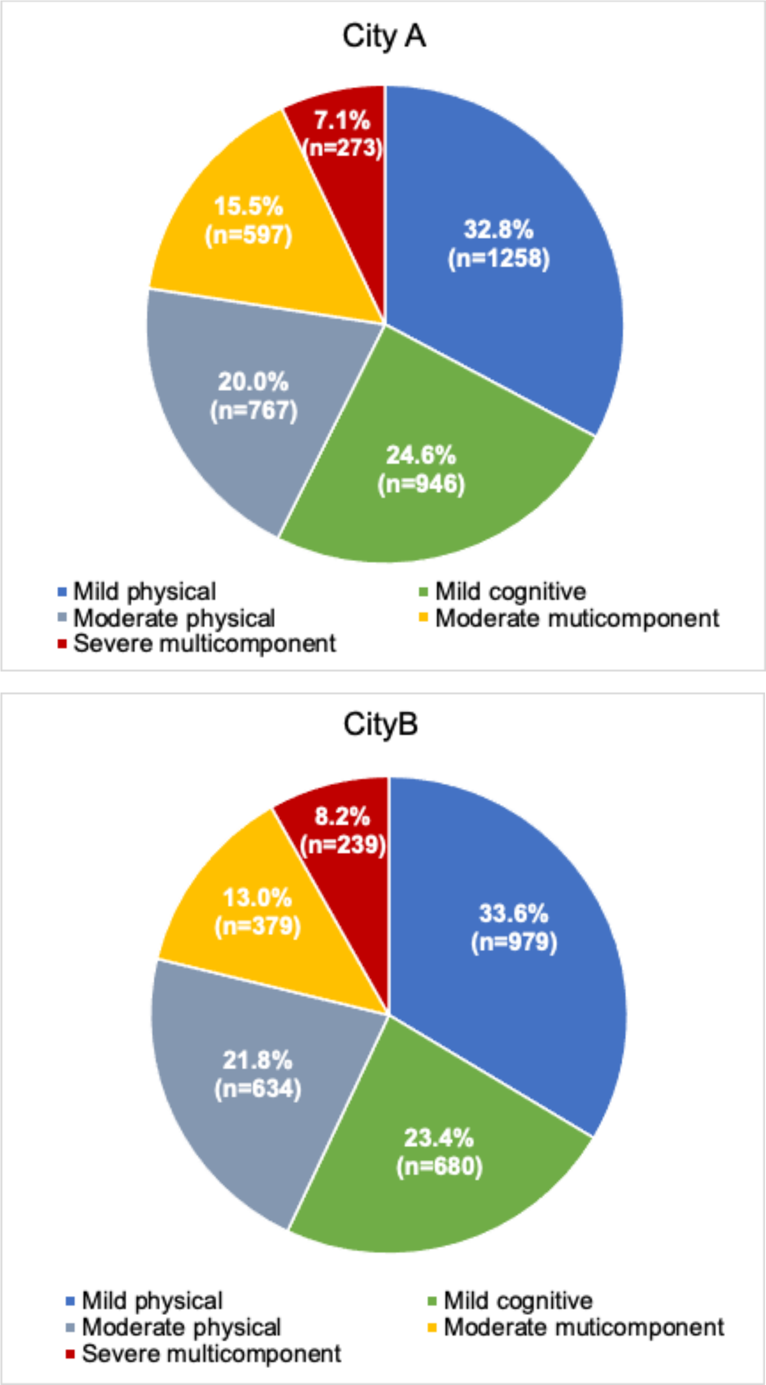


**Figure S4. Proportion of Each Subtype in City A and City B for the Overall Population**

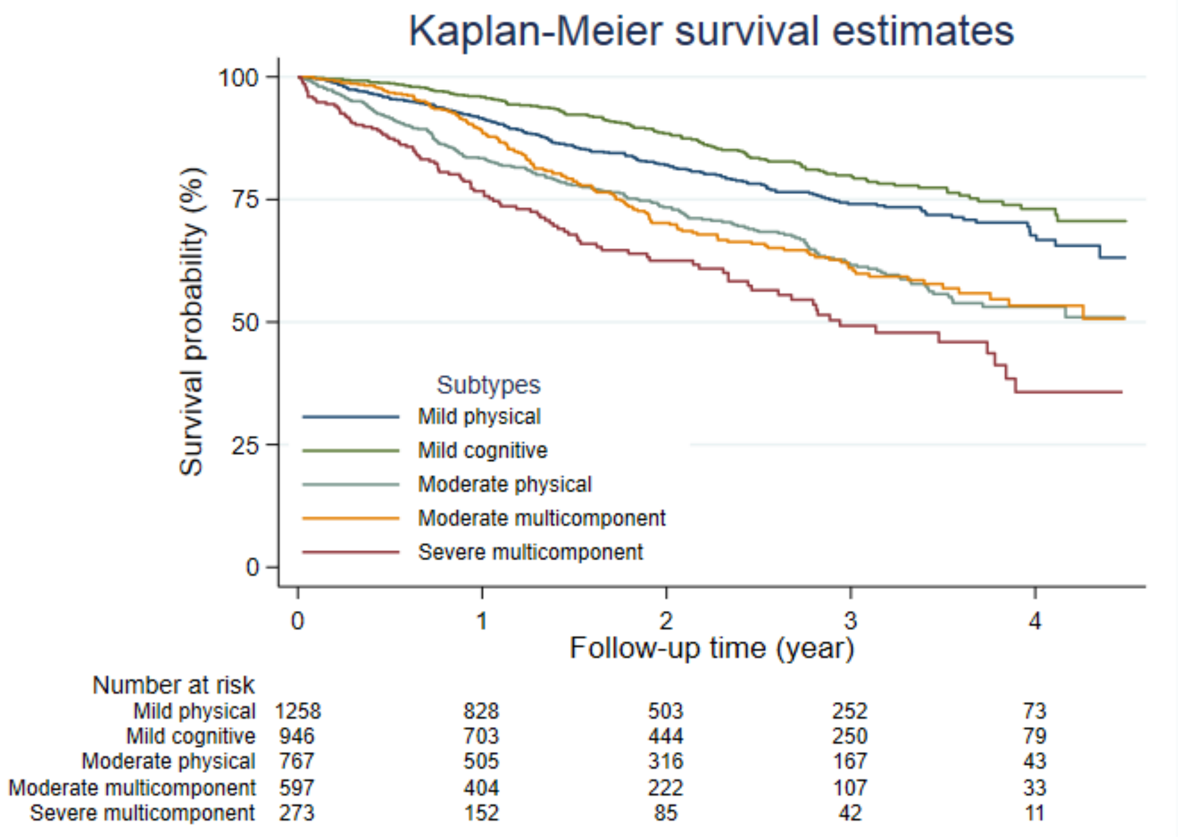


**Figure S5. Kaplan–Meier Survival Curves for Each Subtype**

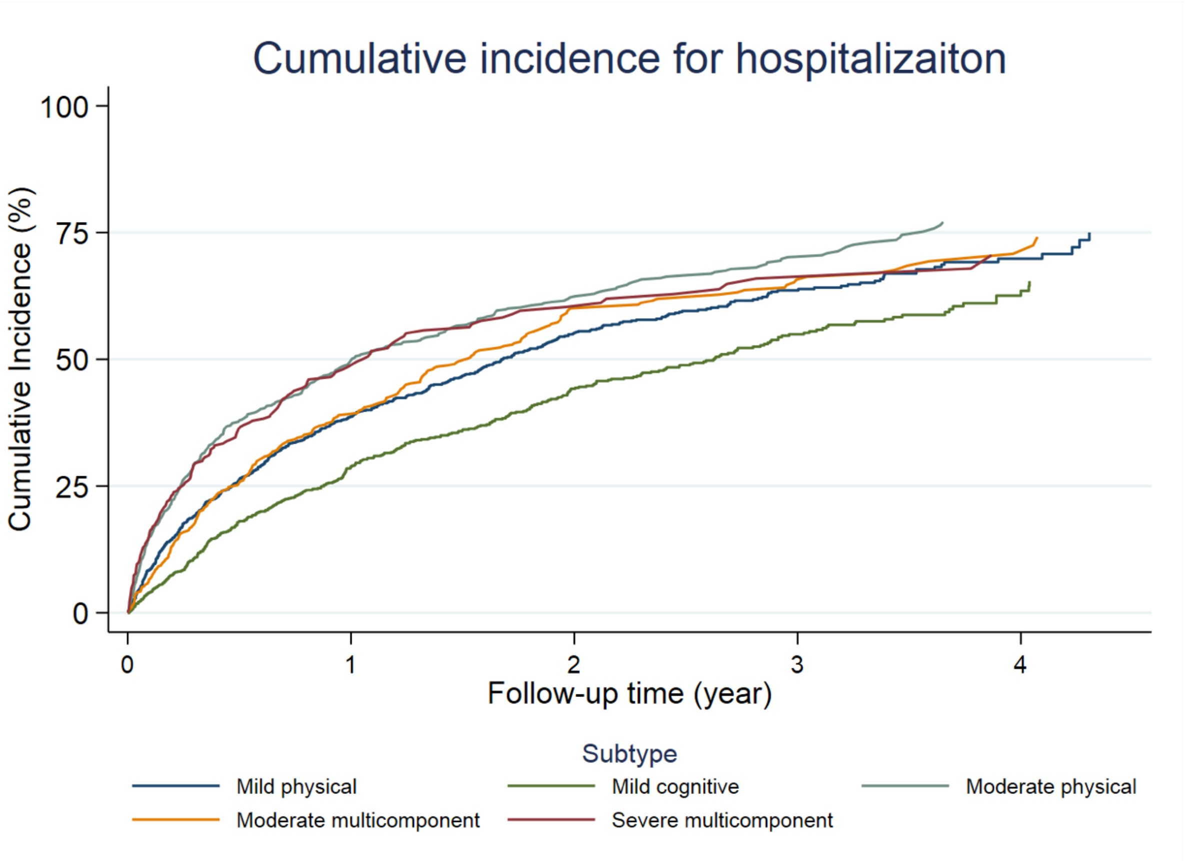


**Figure S6. Cumulative Incidence of Hospital Admission for Each Subtype**

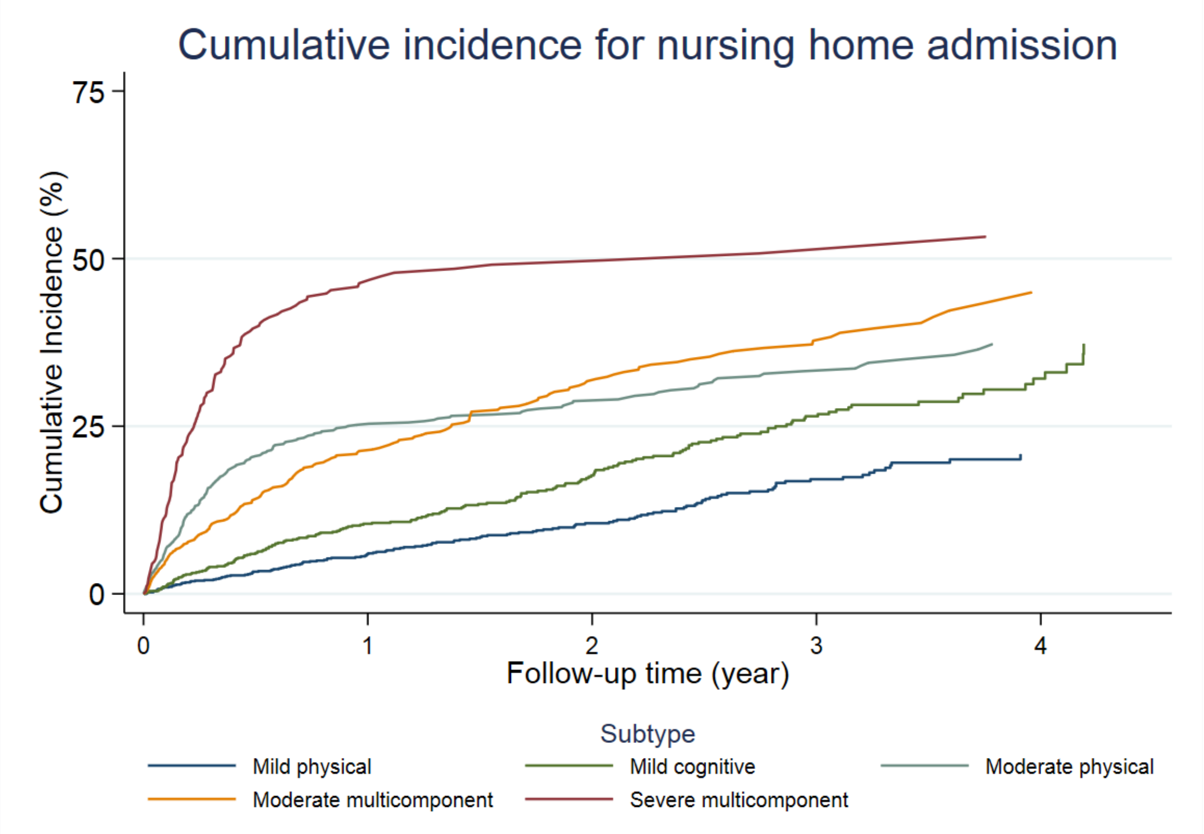


**Figure S7. Cumulative Incidence of Nursing Home Admission for Each Subtype**

**Table S1. Comparisons of the Model Fit Between the Different Numbers of Subtypes**

| Number of subtypes | AIC | BIC | Smallest subtype size (%) | Posterior probability |
|---|---|---|---|---|
| 2 | 193,906.4 | 194,838.1 | 39.4 | 0.98 |
| 3 | 185,171.0 | 186,571.8 | 30.4 | 0.97 |
| 4 | 180,192.7 | 182,062.5 | 15.3 | 0.94 |
| 5 | 176,349.1 | 178,687.9 | 7.1 | 0.94 |
| 6 | 174,867.2 | 177,675.0 | 4.8 | 0.93 |
| 7 | 173583.6 | 176860.5 | 4.5 | 0.93 |
| 8 | 172758.8 | 176504.6 | 4.5 | 0.92 |
| 9 | 172012.0 | 176226.8 | 3.9 | 0.90 |

Abbreviations: AIC, Akaike Information Criterion; BIC, Bayesian Information Criterion

**Table S2. Item Response Probabilities for Physical and Cognitive Functions, Behavioral Problems, and Medical Procedures for Each Subtype in City A**

| | | Subtype 1: mild physical (n = 1258; 32.8 %) | Subtype 2: mild cognitive (n = 946; 24.6%) | Subtype 3: moderate physical (n = 767; 20.0%) | Subtype 4: moderate multicomponent (n = 597; 15.5%) | Subtype 5: severe multicomponent (n = 273; 7.1%) |
|---|---|---|---|---|---|---|
| 1 | Paralysis (left-upper limb) | 0.11 | 0.02 | 0.19 | 0.03 | 0.42 |
| 2 | Paralysis (right-upper limb) | 0.13 | 0.02 | 0.17 | 0.06 | 0.45 |
| 3 | Paralysis (left-lower limb) | 0.35 | 0.07 | 0.53 | 0.17 | 0.69 |
| 4 | Paralysis (right-lower limb) | 0.35 | 0.07 | 0.54 | 0.17 | 0.72 |
| 5 | Paralysis (other) | 0.21 | 0.06 | 0.23 | 0.07 | 0.26 |
| 6 | Contracture (shoulder joint) | 0.11 | 0.01 | 0.13 | 0.04 | 0.21 |
| 7 | Contracture (hip joint) | 0.02 | 0.00 | 0.02 | 0.01 | 0.03 |
| 8 | Contracture (knee) | 0.13 | 0.06 | 0.17 | 0.09 | 0.27 |
| 9 | Contractures (other) | 0.18 | 0.06 | 0.19 | 0.10 | 0.25 |
| 10 | Turning over in bed | 0.13 | 0.05 | 0.22 | 0.08 | 0.58 |
| 11 | Sitting up in bed | 0.05 | 0.00 | 0.43 | 0.03 | 0.77 |
| 12 | Sitting | 0.26 | 0.07 | 0.61 | 0.16 | 0.86 |
| 13 | Both-leg standing | 0.69 | 0.14 | 0.94 | 0.43 | 1.00 |
| 14 | Walking | 0.19 | 0.01 | 0.71 | 0.12 | 0.96 |
| 15 | Standing up | 0.02 | 0.00 | 0.43 | 0.02 | 0.86 |
| 16 | Single leg standing | 0.99 | 0.68 | 1.00 | 0.91 | 1.00 |
| 17 | Washing whole body | 0.84 | 0.26 | 1.00 | 0.87 | 1.00 |
| 18 | Trimming the nails | 0.80 | 0.24 | 0.98 | 0.82 | 1.00 |
| 19 | Eyesight | 0.21 | 0.12 | 0.19 | 0.24 | 0.35 |
| 20 | Hearing ability | 0.55 | 0.49 | 0.51 | 0.54 | 0.57 |
| 21 | Transferring | 0.30 | 0.04 | 0.87 | 0.36 | 0.99 |
| 22 | Moving | 0.50 | 0.13 | 0.96 | 0.63 | 0.99 |
| 23 | Swallowing | 0.14 | 0.09 | 0.14 | 0.14 | 0.52 |
| 24 | Eating | 0.07 | 0.03 | 0.13 | 0.14 | 0.65 |
| 25 | Regulating urination | 0.23 | 0.17 | 0.90 | 0.67 | 1.00 |
| 26 | Regulating defecation | 0.17 | 0.08 | 0.91 | 0.54 | 1.00 |
| 27 | Oral hygiene | 0.27 | 0.07 | 0.91 | 0.66 | 1.00 |
| 28 | Washing face | 0.26 | 0.02 | 0.90 | 0.63 | 1.00 |
| 29 | Combing/Styling hair | 0.18 | 0.02 | 0.76 | 0.57 | 0.96 |
| 30 | Upper-body dressing | 0.45 | 0.08 | 0.94 | 0.84 | 1.00 |
| 31 | Lower-body dressing | 0.51 | 0.07 | 0.99 | 0.85 | 1.00 |
| 32 | Frequency of going out | 0.72 | 0.36 | 0.90 | 0.69 | 0.96 |
| 33 | Taking medicine | 0.75 | 0.84 | 0.95 | 0.99 | 1.00 |
| 34 | Managing money | 0.68 | 0.77 | 0.87 | 0.99 | 0.99 |
| 35 | Daily decision making | 0.03 | 0.24 | 0.10 | 0.74 | 0.87 |
| 36 | Maladaptation to group | 0.00 | 0.01 | 0.00 | 0.02 | 0.03 |
| 37 | Shopping | 0.97 | 0.88 | 1.00 | 0.99 | 1.00 |
| 38 | Cooking | 0.90 | 0.71 | 0.99 | 0.95 | 0.79 |
| 39 | Communicate intentions to others | 0.08 | 0.20 | 0.12 | 0.44 | 0.72 |
| 40 | Understand daily routine | 0.01 | 0.12 | 0.05 | 0.57 | 0.85 |
| 41 | Remember the date of birth | 0.01 | 0.01 | 0.01 | 0.14 | 0.44 |
| 42 | Short-term memory | 0.10 | 0.63 | 0.10 | 0.73 | 0.82 |
| 43 | Remembering own name | 0.00 | 0.00 | 0.00 | 0.02 | 0.21 |
| 44 | Recognizing the season | 0.03 | 0.19 | 0.07 | 0.39 | 0.67 |
| 45 | Location awareness | 0.00 | 0.01 | 0.00 | 0.09 | 0.37 |
| 46 | Wandering | 0.00 | 0.05 | 0.00 | 0.18 | 0.04 |
| 47 | Being lost | 0.00 | 0.05 | 0.01 | 0.17 | 0.03 |

| | | | | | | |
|---|---|---|---|---|---|---|
| 48 | Frequency of feeling persecuted | 0.02 | 0.30 | 0.03 | 0.26 | 0.07 |
| 49 | Making up a story | 0.04 | 0.38 | 0.06 | 0.40 | 0.17 |
| 50 | Emotional instability | 0.10 | 0.37 | 0.07 | 0.34 | 0.24 |
| 51 | Reversal of day and night | 0.01 | 0.10 | 0.02 | 0.19 | 0.16 |
| 52 | Repeating the same story | 0.13 | 0.64 | 0.10 | 0.51 | 0.22 |
| 53 | Shouting | 0.02 | 0.18 | 0.03 | 0.18 | 0.14 |
| 54 | Resisting advice or care | 0.01 | 0.05 | 0.01 | 0.15 | 0.14 |
| 55 | Restlessness | 0.00 | 0.05 | 0.01 | 0.19 | 0.09 |
| 56 | Frequent behavior to go out alone | 0.00 | 0.08 | 0.00 | 0.16 | 0.01 |
| 57 | Collecting items inappropriately | 0.00 | 0.05 | 0.00 | 0.11 | 0.02 |
| 58 | Destruction of things/clothes | 0.00 | 0.03 | 0.00 | 0.04 | 0.02 |
| 59 | Forgetfulness | 0.16 | 0.69 | 0.09 | 0.63 | 0.18 |
| 60 | Monology | 0.03 | 0.16 | 0.02 | 0.25 | 0.20 |
| 61 | Selfish behavior inappropriate for the situation | 0.05 | 0.21 | 0.07 | 0.27 | 0.19 |
| 62 | Incoherent talk | 0.05 | 0.26 | 0.03 | 0.40 | 0.25 |
| 63 | Intravenous infusion | 0.03 | 0.01 | 0.06 | 0.02 | 0.13 |
| 64 | Intravenous hyperalimentation | 0.00 | 0.00 | 0.01 | 0.00 | 0.01 |
| 65 | Dialysis | 0.03 | 0.01 | 0.04 | 0.01 | 0.01 |
| 66 | Care for ostomy | 0.01 | 0.00 | 0.01 | 0.01 | 0.01 |
| 67 | Oxygen therapy | 0.03 | 0.00 | 0.05 | 0.02 | 0.05 |
| 68 | Artificial ventilator | 0.00 | 0.00 | 0.00 | 0.00 | 0.01 |
| 69 | Care for tracheostomy | 0.00 | 0.00 | 0.00 | 0.00 | 0.02 |
| 70 | Pain care | 0.01 | 0.00 | 0.01 | 0.00 | 0.01 |
| 71 | Tube feeding | 0.00 | 0.00 | 0.00 | 0.00 | 0.23 |
| 72 | Monitoring | 0.01 | 0.00 | 0.04 | 0.01 | 0.08 |
| 73 | Pressure ulcer care | 0.01 | 0.00 | 0.04 | 0.00 | 0.11 |
| 74 | Incontinence care | 0.02 | 0.00 | 0.08 | 0.01 | 0.15 |

We highlighted the unique characteristic of each clinical subtype based on an observed/expected ratio ≥ 2 (red characters) and exclusivity ≥ 25% (yellow cells) in line with those in previous studies.

Each variable was converted to a binary form: 1 if the individual had a disability or difficulty requiring assistance from others, or a necessary medical procedure; and 0 otherwise.

**Table S3. Item Response Probabilities of Clusters of Physical and Cognitive Functions, Behavioral Problems, and Medical Procedures for Each Subtype in City B**

| | | Subtype 1: mild physical (n = 979; 33.6 %) | Subtype 2: mild cognitive (n = 680; 23.4%) | Subtype 3: moderate physical (n = 634; 21.8%) | Subtype 4: moderate multicomponent (n = 379; 13.0%) | Subtype 5: severe multicomponent (n = 239; 8.2%) |
|---|---|---|---|---|---|---|
| 1 | Paralysis (left-upper limb) | 0.08 | 0.01 | 0.17 | 0.03 | 0.45 |
| 2 | Paralysis (right-upper limb) | 0.08 | 0.02 | 0.16 | 0.03 | 0.47 |
| 3 | Paralysis (left-lower limb) | 0.43 | 0.10 | 0.64 | 0.23 | 0.79 |
| 4 | Paralysis (right-lower limb) | 0.42 | 0.09 | 0.62 | 0.23 | 0.81 |
| 5 | Paralysis (other) | 0.08 | 0.02 | 0.11 | 0.02 | 0.05 |
| 6 | Contracture (shoulder joint) | 0.09 | 0.02 | 0.12 | 0.03 | 0.14 |
| 7 | Contracture (hip joint) | 0.03 | 0.00 | 0.06 | 0.01 | 0.09 |
| 8 | Contracture (knee) | 0.13 | 0.03 | 0.20 | 0.07 | 0.18 |
| 9 | Contractures (other) | 0.09 | 0.04 | 0.12 | 0.07 | 0.08 |
| 10 | Turning over in bed | 0.06 | 0.02 | 0.30 | 0.05 | 0.61 |
| 11 | Sitting up in bed | 0.07 | 0.01 | 0.63 | 0.04 | 0.86 |
| 12 | Sitting | 0.25 | 0.08 | 0.71 | 0.17 | 0.92 |
| 13 | Both-leg standing | 0.66 | 0.09 | 0.98 | 0.38 | 0.99 |
| 14 | Walking | 0.23 | 0.00 | 0.83 | 0.13 | 0.96 |
| 15 | Standing up | 0.03 | 0.00 | 0.61 | 0.04 | 0.94 |
| 16 | Single leg standing | 0.98 | 0.59 | 1.00 | 0.80 | 1.00 |
| 17 | Washing whole body | 0.88 | 0.31 | 1.00 | 0.91 | 1.00 |
| 18 | Trimming the nails | 0.73 | 0.21 | 0.99 | 0.77 | 1.00 |
| 19 | Eyesight | 0.18 | 0.16 | 0.17 | 0.22 | 0.47 |
| 20 | Hearing ability | 0.38 | 0.37 | 0.36 | 0.42 | 0.54 |
| 21 | Transferring | 0.45 | 0.05 | 0.97 | 0.40 | 1.00 |
| 22 | Moving | 0.65 | 0.17 | 0.98 | 0.58 | 0.99 |
| 23 | Swallowing | 0.15 | 0.10 | 0.23 | 0.12 | 0.60 |
| 24 | Eating | 0.11 | 0.06 | 0.28 | 0.21 | 0.73 |
| 25 | Regulating urination | 0.33 | 0.09 | 0.98 | 0.65 | 1.00 |
| 26 | Regulating defecation | 0.26 | 0.06 | 0.96 | 0.53 | 1.00 |
| 27 | Oral hygiene | 0.27 | 0.06 | 0.93 | 0.79 | 1.00 |
| 28 | Washing face | 0.39 | 0.04 | 0.96 | 0.76 | 1.00 |
| 29 | Combing/Styling hair | 0.15 | 0.02 | 0.69 | 0.69 | 0.97 |
| 30 | Upper-body dressing | 0.48 | 0.11 | 0.96 | 0.83 | 1.00 |
| 31 | Lower-body dressing | 0.59 | 0.11 | 1.00 | 0.84 | 1.00 |
| 32 | Frequency of going out | 0.77 | 0.36 | 0.95 | 0.76 | 0.99 |
| 33 | Taking medicine | 0.74 | 0.73 | 0.95 | 0.98 | 0.96 |
| 34 | Managing money | 0.59 | 0.70 | 0.83 | 0.98 | 1.00 |
| 35 | Daily decision making | 0.01 | 0.06 | 0.07 | 0.54 | 0.87 |
| 36 | Maladaptation to group | 0.00 | 0.01 | 0.00 | 0.03 | 0.02 |
| 37 | Shopping | 0.97 | 0.84 | 1.00 | 0.99 | 0.99 |
| 38 | Cooking | 0.87 | 0.68 | 0.98 | 0.98 | 0.78 |
| 39 | Communicate intentions to others | 0.05 | 0.15 | 0.11 | 0.45 | 0.82 |
| 40 | Understand daily routine | 0.00 | 0.10 | 0.09 | 0.65 | 0.93 |
| 41 | Remember the date of birth | 0.00 | 0.02 | 0.01 | 0.15 | 0.47 |
| 42 | Short-term memory | 0.04 | 0.49 | 0.09 | 0.77 | 0.83 |
| 43 | Remembering own name | 0.00 | 0.00 | 0.00 | 0.02 | 0.22 |
| 44 | Recognizing the season | 0.01 | 0.11 | 0.07 | 0.47 | 0.75 |
| 45 | Location awareness | 0.00 | 0.01 | 0.01 | 0.14 | 0.51 |
| 46 | Wandering | 0.00 | 0.03 | 0.01 | 0.18 | 0.02 |
| 47 | Being lost | 0.00 | 0.04 | 0.00 | 0.12 | 0.00 |
| 48 | Frequency of feeling persecuted | 0.01 | 0.16 | 0.02 | 0.20 | 0.04 |

| | | | | | | |
|---|---|---|---|---|---|---|
| 49 | Making up a story | 0.01 | 0.19 | 0.05 | 0.25 | 0.08 |
| 50 | Emotional instability | 0.08 | 0.24 | 0.09 | 0.29 | 0.15 |
| 51 | Reversal of day and night | 0.03 | 0.07 | 0.06 | 0.26 | 0.08 |
| 52 | Repeating the same story | 0.05 | 0.41 | 0.09 | 0.38 | 0.11 |
| 53 | Shouting | 0.01 | 0.08 | 0.03 | 0.18 | 0.10 |
| 54 | Resisting advice or care | 0.01 | 0.07 | 0.03 | 0.19 | 0.13 |
| 55 | Restlessness | 0.00 | 0.02 | 0.02 | 0.14 | 0.03 |
| 56 | Frequent behavior to go out alone | 0.00 | 0.03 | 0.00 | 0.13 | 0.02 |
| 57 | Collecting items inappropriately | 0.00 | 0.03 | 0.00 | 0.06 | 0.00 |
| 58 | Destruction of things/clothes | 0.00 | 0.01 | 0.00 | 0.03 | 0.01 |
| 59 | Forgetfulness | 0.07 | 0.61 | 0.13 | 0.73 | 0.23 |
| 60 | Monology | 0.00 | 0.05 | 0.01 | 0.18 | 0.09 |
| 61 | Selfish behavior inappropriate for the situation | 0.01 | 0.08 | 0.03 | 0.20 | 0.10 |
| 62 | Incoherent talk | 0.02 | 0.17 | 0.05 | 0.33 | 0.19 |
| 63 | Intravenous infusion | 0.12 | 0.06 | 0.25 | 0.05 | 0.43 |
| 64 | Intravenous hyperalimentation | 0.02 | 0.01 | 0.04 | 0.00 | 0.10 |
| 65 | Dialysis | 0.03 | 0.01 | 0.01 | 0.00 | 0.02 |
| 66 | Care for ostomy | 0.02 | 0.01 | 0.02 | 0.01 | 0.02 |
| 67 | Oxygen therapy | 0.06 | 0.03 | 0.10 | 0.01 | 0.15 |
| 68 | Artificial ventilator | 0.00 | 0.00 | 0.00 | 0.00 | 0.04 |
| 69 | Care for tracheostomy | 0.01 | 0.01 | 0.01 | 0.00 | 0.08 |
| 70 | Pain care | 0.09 | 0.03 | 0.10 | 0.01 | 0.04 |
| 71 | Tube feeding | 0.01 | 0.01 | 0.02 | 0.00 | 0.25 |
| 72 | Monitoring | 0.04 | 0.01 | 0.08 | 0.02 | 0.18 |
| 73 | Pressure ulcer care | 0.01 | 0.00 | 0.05 | 0.02 | 0.08 |
| 74 | Incontinence care | 0.04 | 0.01 | 0.18 | 0.03 | 0.35 |

We highlighted the unique characteristic of each clinical subtype based on an observed/expected ratio ≥ 2 (red characters) and exclusivity ≥ 25% (yellow cells) in line with those in previous studies.

Each variable was converted to a binary form: 1 if the individual had a disability or difficulty requiring assistance from others, or a necessary medical procedure; and 0 otherwise.

**Table S4. Baseline Characteristics by Physical and Cognitive Functions, Behavioral Problems, and Medical Procedures Subtypes in City B**

| | Subtype 1: mild physical (n = 979; 33.6 %) | Subtype 2: mild cognitive (n = 680; 23.4%) | Subtype 3: moderate physical (n = 634; 21.8%) | Subtype 4: moderate multicomponent (n = 379; 13.0%) | Subtype 5: severe multicomponent (n = 239; 8.2%) | Overall (n = 2911) |
|---|---|---|---|---|---|---|
| Age, years, median (IQR) | 82 (77–87) | 82 (77–87) | 82 (77–87) | 82 (77–87) | 82 (72–87) | 82 (77–87) |
| Age category, years | | | | | | |
| 65–74 | 231 (23.6%) | 115 (16.9%) | 151 (23.8%) | 59 (15.6%) | 61 (25.5%) | 617 (21.2%) |
| 75–84 | 432 (44.1%) | 328 (48.2%) | 272 (42.9%) | 150 (39.6%) | 103 (43.1%) | 1285 (44.1%) |
| 85–94 | 294 (30.0%) | 226 (33.2%) | 189 (29.8%) | 156 (41.2%) | 65 (27.2%) | 930 (31.9%) |
| ≥ 95 | 22 (2.2%) | 11 (1.6%) | 22 (3.5%) | 14 (3.7%) | 10 (4.2%) | 79 (2.7%) |
| Sex, female | 532 (54.3%) | 402 (59.1%) | 316 (49.8%) | 184 (48.5%) | 117 (49.0%) | 1551 (53.3%) |
| Care need level | | | | | | |
| 1 | 487 (49.7%) | 538 (79.1%) | 9 (1.4%) | 95 (25.1%) | 0 (0.0%) | 1129 (38.8%) |
| 2 | 398 (40.7%) | 112 (16.5%) | 95 (15.0%) | 171 (45.1%) | 1 (0.4%) | 777 (26.7%) |
| 3 | 61 (6.2%) | 14 (2.1%) | 195 (30.8%) | 79 (20.8%) | 27 (11.3%) | 376 (12.9%) |
| 4 | 23 (2.3%) | 16 (2.4%) | 242 (38.2%) | 32 (8.4%) | 75 (31.4%) | 388 (13.3%) |
| 5 | 10 (1.0%) | 0 (0.0%) | 93 (14.7%) | 2 (0.5%) | 136 (56.9%) | 241 (8.3%) |

Abbreviation: CI, confidence interval; IQR, interquartile range.

Data were reported as numbers (%) for categorical variables and as medians (IQR) for continuous variables.

**Table S5. Results of Additional Analysis Including Underlying Diseases for Clustering in City A**

| | | Subtype 1: mild physical (n = 1284; 33.4 %) | Subtype 2: mild cognitive (n = 901; 23.5%) | Subtype 3: moderate physical (n = 789; 20.5%) | Subtype 4: moderate multicomponent (n = 582; 15.2%) | Subtype 5: severe multicomponent (n = 285; 7.4%) |
|---|---|---|---|---|---|---|
| 1 | Paralysis (left-upper limb) | 0.11 | 0.02 | 0.19 | 0.04 | 0.39 |
| 2 | Paralysis (right-upper limb) | 0.13 | 0.01 | 0.17 | 0.07 | 0.43 |
| 3 | Paralysis (left-lower limb) | 0.35 | 0.07 | 0.52 | 0.18 | 0.67 |
| 4 | Paralysis (right-lower limb) | 0.34 | 0.07 | 0.53 | 0.17 | 0.71 |
| 5 | Paralysis (other) | 0.21 | 0.06 | 0.23 | 0.07 | 0.25 |
| 6 | Contracture (shoulder joint) | 0.11 | 0.01 | 0.12 | 0.04 | 0.20 |
| 7 | Contracture (hip joint) | 0.02 | 0.00 | 0.02 | 0.01 | 0.03 |
| 8 | Contracture (knee) | 0.13 | 0.05 | 0.17 | 0.09 | 0.27 |
| 9 | Contractures (other) | 0.18 | 0.06 | 0.19 | 0.10 | 0.25 |
| 10 | Turning over in bed | 0.13 | 0.05 | 0.22 | 0.08 | 0.56 |
| 11 | Sitting up in bed | 0.05 | 0.00 | 0.42 | 0.02 | 0.77 |
| 12 | Sitting | 0.25 | 0.07 | 0.60 | 0.16 | 0.85 |
| 13 | Both-leg standing | 0.67 | 0.13 | 0.94 | 0.42 | 0.99 |
| 14 | Walking | 0.18 | 0.01 | 0.69 | 0.12 | 0.95 |
| 15 | Standing up | 0.01 | 0.00 | 0.41 | 0.02 | 0.85 |
| 16 | Single leg standing | 0.99 | 0.68 | 1.00 | 0.91 | 1.00 |
| 17 | Washing whole body | 0.82 | 0.26 | 1.00 | 0.88 | 1.00 |
| 18 | Trimming the nails | 0.78 | 0.23 | 0.98 | 0.82 | 1.00 |
| 19 | Eyesight | 0.20 | 0.12 | 0.19 | 0.23 | 0.34 |
| 20 | Hearing ability | 0.55 | 0.49 | 0.52 | 0.54 | 0.56 |
| 21 | Transferring | 0.28 | 0.04 | 0.87 | 0.35 | 0.99 |
| 22 | Moving | 0.49 | 0.13 | 0.96 | 0.63 | 0.99 |
| 23 | Swallowing | 0.14 | 0.10 | 0.14 | 0.14 | 0.52 |
| 24 | Eating | 0.07 | 0.04 | 0.12 | 0.15 | 0.64 |
| 25 | Regulating urination | 0.23 | 0.18 | 0.88 | 0.67 | 1.00 |
| 26 | Regulating defecation | 0.16 | 0.09 | 0.89 | 0.55 | 1.00 |
| 27 | Oral hygiene | 0.25 | 0.07 | 0.90 | 0.68 | 1.00 |
| 28 | Washing face | 0.24 | 0.02 | 0.90 | 0.63 | 1.00 |
| 29 | Combing/Styling hair | 0.16 | 0.02 | 0.74 | 0.58 | 0.96 |
| 30 | Upper-body dressing | 0.43 | 0.08 | 0.94 | 0.84 | 1.00 |
| 31 | Lower-body dressing | 0.49 | 0.07 | 0.99 | 0.86 | 1.00 |
| 32 | Frequency of going out | 0.71 | 0.36 | 0.89 | 0.69 | 0.96 |
| 33 | Taking medicine | 0.74 | 0.85 | 0.95 | 0.99 | 1.00 |
| 34 | Managing money | 0.67 | 0.78 | 0.86 | 0.99 | 0.99 |
| 35 | Daily decision making | 0.02 | 0.25 | 0.09 | 0.73 | 0.87 |
| 36 | Maladaptation to group | 0.00 | 0.01 | 0.00 | 0.02 | 0.03 |
| 37 | Shopping | 0.97 | 0.88 | 1.00 | 0.99 | 1.00 |
| 38 | Cooking | 0.90 | 0.72 | 0.99 | 0.95 | 0.80 |
| 39 | Communicate intentions to others | 0.08 | 0.20 | 0.12 | 0.43 | 0.72 |
| 40 | Understand daily routine | 0.01 | 0.13 | 0.05 | 0.57 | 0.83 |
| 41 | Remember the date of birth | 0.01 | 0.02 | 0.00 | 0.15 | 0.44 |
| 42 | Short-term memory | 0.11 | 0.63 | 0.10 | 0.74 | 0.81 |
| 43 | Remembering own name | 0.00 | 0.00 | 0.00 | 0.02 | 0.20 |
| 44 | Recognizing the season | 0.03 | 0.20 | 0.07 | 0.40 | 0.66 |
| 45 | Location awareness | 0.00 | 0.01 | 0.00 | 0.09 | 0.35 |
| 46 | Wandering | 0.00 | 0.05 | 0.00 | 0.18 | 0.04 |
| 47 | Being lost | 0.00 | 0.05 | 0.01 | 0.17 | 0.03 |
| 48 | Frequency of feeling persecuted | 0.03 | 0.30 | 0.03 | 0.25 | 0.06 |

| | | | | | | |
|---|---|---|---|---|---|---|
| 49 | Making up a story | 0.04 | 0.39 | 0.06 | 0.39 | 0.17 |
| 50 | Emotional instability | 0.10 | 0.38 | 0.07 | 0.34 | 0.23 |
| 51 | Reversal of day and night | 0.01 | 0.10 | 0.02 | 0.19 | 0.16 |
| 52 | Repeating the same story | 0.14 | 0.65 | 0.10 | 0.50 | 0.21 |
| 53 | Shouting | 0.03 | 0.18 | 0.03 | 0.18 | 0.14 |
| 54 | Resisting advice or care | 0.01 | 0.05 | 0.01 | 0.15 | 0.13 |
| 55 | Restlessness | 0.00 | 0.05 | 0.01 | 0.19 | 0.10 |
| 56 | Frequent behavior to go out alone | 0.00 | 0.09 | 0.00 | 0.16 | 0.01 |
| 57 | Collecting items inappropriately | 0.00 | 0.05 | 0.00 | 0.10 | 0.02 |
| 58 | Destruction of things/clothes | 0.00 | 0.04 | 0.00 | 0.04 | 0.02 |
| 59 | Forgetfulness | 0.17 | 0.70 | 0.08 | 0.63 | 0.18 |
| 60 | Monology | 0.03 | 0.17 | 0.02 | 0.25 | 0.20 |
| 61 | Selfish behavior inappropriate for the situation | 0.05 | 0.22 | 0.08 | 0.27 | 0.19 |
| 62 | Incoherent talk | 0.05 | 0.27 | 0.03 | 0.39 | 0.24 |
| 63 | Intravenous infusion | 0.03 | 0.01 | 0.06 | 0.02 | 0.12 |
| 64 | Intravenous hyperalimentation | 0.00 | 0.00 | 0.00 | 0.00 | 0.01 |
| 65 | Dialysis | 0.03 | 0.01 | 0.04 | 0.01 | 0.01 |
| 66 | Care for ostomy | 0.01 | 0.00 | 0.01 | 0.01 | 0.01 |
| 67 | Oxygen therapy | 0.03 | 0.00 | 0.05 | 0.02 | 0.05 |
| 68 | Artificial ventilator | 0.00 | 0.00 | 0.00 | 0.00 | 0.01 |
| 69 | Care for tracheostomy | 0.00 | 0.00 | 0.00 | 0.00 | 0.02 |
| 70 | Pain care | 0.01 | 0.00 | 0.01 | 0.00 | 0.01 |
| 71 | Tube feeding | 0.00 | 0.00 | 0.00 | 0.00 | 0.22 |
| 72 | Monitoring | 0.01 | 0.00 | 0.04 | 0.01 | 0.07 |
| 73 | Pressure ulcer care | 0.01 | 0.00 | 0.04 | 0.00 | 0.10 |
| 74 | Incontinence care | 0.02 | 0.00 | 0.08 | 0.01 | 0.15 |
| 75 | Hemorrhagic stroke | 0.04 | 0.02 | 0.06 | 0.04 | 0.18 |
| 76 | Ischemic stroke | 0.25 | 0.32 | 0.29 | 0.32 | 0.33 |
| 77 | Other cerebrovascular diseases | 0.23 | 0.17 | 0.24 | 0.21 | 0.32 |
| 78 | Ischemic heart disease | 0.34 | 0.23 | 0.33 | 0.23 | 0.22 |
| 79 | Arrhythmia | 0.26 | 0.23 | 0.29 | 0.23 | 0.35 |
| 80 | Heart failure | 0.29 | 0.16 | 0.35 | 0.23 | 0.29 |
| 81 | Other cardiac diseases | 0.12 | 0.08 | 0.16 | 0.08 | 0.10 |
| 82 | Cancer | 0.23 | 0.15 | 0.24 | 0.15 | 0.20 |
| 83 | Chronic obstructive pulmonary disease | 0.08 | 0.04 | 0.08 | 0.07 | 0.05 |
| 84 | Pneumonia | 0.10 | 0.03 | 0.22 | 0.11 | 0.30 |
| 85 | Other lower respiratory tract disease | 0.41 | 0.28 | 0.39 | 0.28 | 0.38 |
| 86 | Rheumatoid arthritis | 0.06 | 0.02 | 0.05 | 0.02 | 0.02 |
| 87 | Other arthropathies | 0.47 | 0.34 | 0.48 | 0.33 | 0.25 |
| 88 | Dorsopathies | 0.72 | 0.52 | 0.69 | 0.49 | 0.46 |
| 89 | Dementia | 0.07 | 0.58 | 0.09 | 0.48 | 0.28 |
| 90 | Parkinson’s disease | 0.06 | 0.04 | 0.04 | 0.05 | 0.04 |
| 91 | Insulin-dependent diabetes | 0.10 | 0.02 | 0.14 | 0.06 | 0.16 |
| 92 | Non-insulin-dependent diabetes | 0.32 | 0.29 | 0.31 | 0.34 | 0.23 |
| 93 | Visual impairment | 0.08 | 0.06 | 0.07 | 0.04 | 0.04 |
| 94 | Hearing impairment | 0.05 | 0.04 | 0.04 | 0.03 | 0.04 |
| 95 | Femur fractures | 0.08 | 0.01 | 0.09 | 0.06 | 0.05 |
| 96 | Other fractures | 0.21 | 0.08 | 0.29 | 0.16 | 0.16 |

We highlighted the unique characteristic of each clinical subtype based on an observed/expected ratio ≥ 2 (red characters) and exclusivity ≥ 25% (yellow cells) in line with those in previous studies.

Each variable was converted to a binary form: 1 if the individual had a disability or difficulty requiring assistance from others, or a necessary medical procedure; and 0 otherwise.

**Table S6. Details of the Longitudinal Analysis and Results of the Cox Regression Analysis for Death, Hospitalization, and Nursing Home Admission**

| | Subtype 1: mild physical (n = 1258; 32.8 %) | | Subtype 2: mild cognitive (n = 946; 24.6%) | | Subtype 3: moderate physical (n = 767; 20.0%) | | Subtype 4: moderate multicomponent (n = 597; 15.5%) | | Subtype 5: severe multicomponent (n = 273; 7.1%) | |
|---|---|---|---|---|---|---|---|---|---|---|
| **Death** | | | | | | | | | | |
| Death, per 100 person-year (95%CI) | 9.7 | (8.5–11.0) | 6.8 | (5.7–8.1) | 16.2 | (14.2–18.4) | 15.3 | (13.1–17.8) | 24.4 | (20.1–29.7) |
| Unadjusted hazard ratio (95%CI) | 1 | (reference) | 0.70 | (0.56–0.87) | 1.67 | (1.39–2.02) | 1.58 | (1.29–1.94) | 2.55 | (2.01–3.22) |
| Adjusted hazard ratio (95%CI) † | 1 | (reference) | 0.72 | (0.58–0.89) | 1.57 | (1.31–1.90) | 1.50 | (1.22–1.84) | 2.56 | (2.02–3.24) |
| **Hospitalization** | | | | | | | | | | |
| Hospitalization, per 100 person-year (95%CI) | 42.8 | (39.5–46.2) | 29.0 | (26.3–31.9) | 58.2 | (53.2–63.8) | 45.0 | (40.3–50.3) | 64.3 | (54.8–75.4) |
| Unadjusted hazard ratio (95%CI) | 1 | (reference) | 0.73 | (0.65–0.82) | 1.32 | (1.17–1.50) | 1.04 | (0.91–1.19) | 1.23 | (1.02–1.49) |
| Adjusted hazard ratio (95%CI) † | 1 | (reference) | 0.74 | (0.65–0.83) | 1.32 | (1.16–1.49) | 1.02 | (0.90–1.17) | 1.23 | (1.02–1.48) |
| | Subtype 1: mild physical (n = 1252; 32.9 %) | | Subtype 2: mild cognitive (n = 943; 24.8%) | | Subtype 3: moderate physical (n = 754; 19.8%) | | Subtype 4: moderate multicomponent (n = 591; 15.5%) | | Subtype 5: severe multicomponent (n = 269; 7.1%) | |
| **Nursing home admission** * | | | | | | | | | | |
| Nursing home admission per 100 person-year (95%CI) | 6.7 | (5.7–7.9) | 11.0 | (9.5–12.7) | 21.6 | (18.9–24.7) | 22.0 | (18.9–25.4) | 58.2 | (48.9–69.3) |
| Unadjusted hazard ratio (95%CI) | 1 | (reference) | 1.72 | (1.39–2.13) | 2.88 | (2.33–3.57) | 2.98 | (2.40–3.70) | 5.90 | (4.58–7.60) |
| Adjusted hazard ratio (95%CI) † | 1 | (reference) | 1.68 | (1.35–2.08) | 2.87 | (2.32–3.55) | 2.97 | (2.38–3.69) | 5.91 | (4.57–7.63) |

Abbreviations: CI, confidence interval.

* Those admitted to nursing homes at baseline were excluded.

† Adjusted for age and sex.

**Table S7. Details of Changes in Care-Need Levels and Results of Logistic Regression Analysis for Care-Need Level Deterioration within 2 Years**

| | Subtype 1: mild physical (n = 636; 31.0 %) | Subtype 2: mild cognitive (n = 517; 25.2%) | Subtype 3: moderate physical (n = 433; 21.1%) | Subtype 4: moderate multicomponent (n = 319; 15.5%) | Subtype 5: severe multicomponent (n = 148; 7.2%) |
|---|---|---|---|---|---|
| Improved, n (%) | 26 (4.1) | 8 (1.6) | 108 (24.9) | 32 (10.0) | 25 (16.9) |
| No change, n (%) | 195 (30.7) | 197 (38.1) | 76 (17.6) | 66 (20.7) | 27 (18.2) |
| Worsened, n (%) | 137 (21.5) | 154 (29.8) | 52 (12.0) | 78 (24.5) | 9 (6.1) |
| Dead, n (%) | 178 (28.0) | 115 (22.2) | 163 (37.6) | 123 (38.6) | 79 (53.4) |
| Alive but care-need levels not reassessed within 2 years, n (%) | 93 (14.6) | 33 (6.4) | 25 (5.8) | 18 (5.6) | 5 (3.4) |
| Loss-to-follow-up at 2 years, n (%) | 7 (1.1) | 10 (1.9) | 9 (2.1) | 2 (0.6) | 3 (2.0) |
| Logistic regression analysis for care-need level deterioration* | | | | | |
| Unadjusted odds ratio (95%CI) | 1(reference) | 1.13 (0.89–1.42) | 1.03 (0.80–1.31) | 1.73 (1.31–2.28) | 1.54 (1.07–2.22) |
| Adjusted odds ratio (95%CI) † | 1(reference) | 1.17 (0.92–1.48) | 0.97 (0.75–1.25) | 1.67 (1.26–2.22) | 1.44 (0.99–2.10) |
| Sensitivity analysis‡ | | | | | |
| Unadjusted odds ratio (95%CI) | 1(reference) | 0.92 (0.72–1.18) | 0.82 (0.63–1.06) | 1.44 (1.07–1.94) | 1.19 (0.81–1.74) |
| Adjusted odds ratio (95%CI) † | 1(reference) | 0.97 (0.76–1.26) | 0.80 (0.61–1.04) | 1.44 (1.06–1.94) | 1.12 (0.76–1.66) |

Abbreviation: CI, confidence interval.

* The outcome was worsened care-need levels or death within 2 years.

† Adjusted for age and sex.

‡ Those lost to follow-up, and those alive without reassessment of care-need levels within 2 years were excluded.

**Table S8. Subgroup Analysis by Care-Need Level: Results of the Cox Regression Analysis for Death, Hospitalization, and Nursing Home Admission**

| **Care-need level 1-2 (n=2787)** | Subtype 1: mild physical (n = 1229) | Subtype 2: Behavioral (n = 944) | Subtype 3: moderate physical (n = 213) | Subtype 4: moderate multicomponent (n = 401) | Subtype 5: severe multicomponent |
|---|---|---|---|---|---|
| Adjusted hazard ratio for death (95%CI)† | 1 (reference) | 0.76 (0.61–0.95) | 1.22 (0.90–1.66) | 1.31 (1.02–1.67) | N.A. |
| Adjusted hazard ratio for hospitalization (95%CI)† | 1 (reference) | 0.71 (0.63–0.81) | 1.44 (1.18–1.77) | 0.91 (0.78–1.07) | N.A. |
| Adjusted hazard ratio for nursing home admission (95%CI)*† | 1 (reference) | 2.56 (1.78–3.67) | 4.90 (3.00–8.00) | 3.36 (2.19–5.14) | N.A. |
| **Care-need level 3-5 (n=1019)** | Subtype 1: mild physical | Subtype 2: Behavioral | Subtype 3: moderate physical (n = 554) | Subtype 4: moderate multicomponent (n = 196) | Subtype 5: severe multicomponent (n=269) |
| Adjusted hazard ratio for death (95%CI)† | N.A. | N.A. | 1 (reference) | 1.22 (0.92–1.62) | 1.42 (1.11–1.82) |
| Adjusted hazard ratio for hospitalization (95%CI)† | N.A. | N.A. | 1 (reference) | 0.88 (0.70-1.11) | 0.95 (0.76–1.20) |
| Adjusted hazard ratio for nursing home admission (95%CI)*† | N.A. | N.A. | 1 (reference) | 0.88 (0.70-1.11) | 0.95 (0.76–1.20) |

Abbreviations: CI, confidence interval l; N.A., not available.

* Those admitted to nursing homes at baseline were excluded.

†Adjusted for age and sex.

In the analysis of the care-need level 1–2 subgroup, we excluded subtype 5 due to the small number of the participants. In the analysis of the care-need level 3–5 subgroup, we excluded subtypes 1 and 2 due to the small number of the participants.
